# New Eye Tracking Metrics System: The Value in Early Diagnosis of Autism Spectrum Disorder

**DOI:** 10.1101/2024.08.28.24312596

**Authors:** Raymond Kong Wang, Kenneth Kwong, Kevin Liu, Xue-Jun Kong

## Abstract

**Background:** Eye tracking (ET) is emerging as a promising early and objective screening method for autism spectrum disorders (ASD), but it requires more reliable metrics with enhanced sensitivity and specificity for clinical use.

**Methods:** This study introduces a suite of novel ET metrics: Area of Interest (AOI) Switch Counts (ASC), Favorable AOI Shifts (FAS) along self-determined pathways, and AOI Vacancy Counts (AVC). These metrics were applied to toddlers and preschoolers diagnosed with ASD. The correlation between these new ET metrics and Autism Diagnostic Observation Schedule, Second Edition (ADOS-2) scores was assessed using linear regression. Sensitivity and specificity of the cut-off scores were also evaluated to predict diagnosis.

**Results:** Our findings indicate significantly lower FAS and ASC and higher AVC (P<0.05) in children with ASD compared to their non-ASD counterparts within this high-risk cohort. There were no significant differences in total fixation time or pupil size (p > 0.05). Additionally, FAS was negatively correlated with ADOS-2 total scores and the social affect (SA) subscale (p < 0.05). Among these new ET metrics, AVC yielded the best sensitivity (88-100%) and specificity (80-88%) with a cut-off score of 0.305-0.306, followed by FAS and ASC for distinguishing ASD from non-ASD for diagnosis.

**Conclusions:** This study confirms the utility of innovative ET metrics—FAS, AVC, and ASC, which exhibit markedly improved sensitivity and specificity, enhancing ASD screening and diagnostic processes.

## Introduction

Early diagnosis and intervention are pivotal in determining long-term outcomes for individuals with Autism Spectrum Disorder (ASD). The prognostic implications underscore the necessity for developing readily accessible and effective early detection tools (Towle et al., 2016; Kantzer et al., 2016; Dow et al., 2017; Zwaigenbaum et al., 2018). Current diagnostic frameworks, such as the Diagnostic and Statistical Manual of Mental Disorders, Fifth Edition (DSM-5) (American Psychiatric Association, 2013), and the Autism Diagnostic Observation Schedule, Second Edition (ADOS-2) (Lord et al., 2012), provide structured criteria for early diagnosis. Nevertheless, these conventional methods are often elaborate, time-consuming, and resource-intensive. This complexity can delay diagnostic and therapeutic interventions, particularly in underserved populations (Rotholz et al., 2017; Monteiro et al., 2016). Despite recent advances in diagnostic methodologies, the mean age of diagnosis persists at four to five years (Zwaigenbaum et al., 2018). Furthermore, disparities in diagnosis times are evident, with ethno-racial minorities and non-English speaking children diagnosed significantly later than their white counterparts (Stahmer et al., 2019). Addressing these disparities is critical, underscoring the urgent need for innovative diagnostic tools that facilitate earlier detection and intervention in high-risk populations.

Amidst the subjective limitations of standard assessments, recent research has shifted towards objective biomarkers for ASD diagnosis. Eye tracking (ET) technology has gained prominence as a promising diagnostic tool due to its inherent objectivity and rapid assessment capabilities (Gliga et al., 2015; Kong et al., 2017; Helminen et al., 2017; Wan et al., 2019; Kong et al., 2022). Traditionally utilized in human perception studies and extensively in ASD research, ET technology quantifies eye positions, movements (Helminen et al., 2017; Black et al., 2017), and pupil size dynamics (Nystrom et al., 2018; Lynch et al., 2018; Artoni et al., 2020) to delineate zones of user interest. Distinctive eye movement patterns and gaze behaviors in ASD, such as challenges in interpreting gaze cues, a preference for systematically arranged images over faces, and a lack of right hemispheric dominance for facial processing, have been well-documented (Frazier et al., 2018; Fujioka et al., 2016; Papagiannopoulou et al., 2014; Strathearn et al., 2017). Essential metrics in ASD ET research include Total Gaze Count (TGC) and Total Fixation Time (TFT), which respectively measure the frequency of gazes and the duration of eye fixation within designated Areas of Interest (AOIs) (Wan et al., 2019; Kong et al., 2017; Helminen et al., 2017; Kong et al., 2022). Recent investigations have highlighted significant reductions in TFT across most AOIs for ASD subjects compared to controls in developmental cohorts, reinforcing the diagnostic potential of these parameters (Kong et al., 2022).

Despite the promising aspects of ET, variability in its protocols and paradigms has historically limited its utility as a consistent diagnostic tool (Mastergeorge et al., 2021). In response, this study introduces novel ET metrics such as Area of Interest Switch Counts (ASC), Favored Area of Interest Shifts (FAS), and Area of Interest Vacancy Counts (AVC). These metrics are designed to quantify dynamic shifts between AOIs and are hypothesized to reflect fundamental ASD-related deficits such as joint attention, social referencing, and theory of mind. Integrating these new metrics with established diagnostic assessments like ADOS-2, we aim to significantly enhance the specificity and sensitivity of ET for ASD diagnosis. Preliminary findings have facilitated the identification of optimal cutoff scores, providing foundational proof-of-concept and methodologies poised to refine ASD diagnostic approaches through advanced ET metrics.

## Methods

### Participants

This study involved thirty-nine individuals aged 18 to 84 months, identified as high-risk for Autism Spectrum Disorder (ASD) by clinicians or caregivers in Massachusetts and its surrounding states. High-risk status was confirmed via telephone screening prior to enrollment. Inclusion criteria required participants to meet one or more of the following:

1. having at least one sibling with a clinical ASD diagnosis;
2. caregiver or clinician concerns regarding the child’s development in social interaction, play, or other behaviors;
3. scoring in the positive range on the Modified Checklist for Autism in Toddlers (M-CHAT).

Participants with major congenital or genetic disorders, or behavioral issues likely to cause significant stress during testing were excluded. Those previously diagnosed with ASD were included without disclosing their diagnosis to the examiner. Subjects were categorized into ASD and non-ASD groups based on DSM-5 criteria, evaluated by two field experts.

### Assessment Instruments and Protocols

The Institutional Review Board (IRB) of Massachusetts General Hospital approved this study (MGH, 2017P0000857). Informed consent was obtained from all participants’ legal guardians.

### Eye Tracking Setup

Eye tracking data were collected using a Tobii X3-120 eye tracker, with the screen resolution set to 1024 × 768 pixels, a sampling frequency of 250 Hz, and a spatial resolution of 0.03 degrees. Participants were seated in a dark, soundproof room, 65 cm from a 22-inch widescreen LCD monitor, with their vision centered on the display. Data inclusion required successful completion of a five-point calibration and the full experiment, retaining only data from compliant participants.

### Stimuli

Eye tracking stimuli consisted of two videos previously used in related research (Wan 2019; Kong 2022). The first video (25 seconds) featured a woman and a tablet, alternating attention between the two based on the tablet’s activity (turning on/off), testing joint attention capabilities. The second video (10 seconds) displayed a woman silently mouthing the alphabet, focusing on the eyes and mouth as separate areas of interest (AOIs) to examine social communication and early language development cues (Artoni 2020).

### Autism Diagnostic Observation Schedule (ADOS-2)

ADOS-2, the gold standard in ASD diagnostics, involves several modules selected based on the participant’s age and language development, assessing social interactions, communication, and behaviors (Lord et al., 2016; Falkmer et al., 2013; Luyster et al., 2009). It ends with a diagnostic algorithm tailored to maximize diagnostic sensitivity and specificity. Each module’s outcomes are quantified into a calibrated severity score (CSS) from 1 to 10 (Hus et al., 2014). Administration involved two trained professionals for ADOS-2 and three for eye tracking, with the entire evaluation process taking approximately one hour.

### Statistical Analysis

Eye tracking raw data was processed using Tobii Pro software. Consistent with prior studies (Chawarska 2016; Wang 2018), data segments with less than 25% screen-looking time were excluded, as were participants with fewer than 50% valid trials (Pierce 2011). Comparisons of TGC, ASC, FAS, and AVC between ASD and non-ASD groups utilized the Wilcoxon rank-sum test, while discriminant analysis evaluated the ability of AOIs to categorize subjects by ASD severity. Detailed AOI shift analysis within and across different attention time blocks for video 1 was performed. Correlations between TGC, ASC, FAS, AVC, and ADOS-2 total/sub-scores were examined using R/R-Studio to determine the sensitivity and specificity of ET metrics in predicting ASD diagnosis, identifying optimal cutoff scores for effective separation of ASD and non-ASD groups.

## Results

### Participant Demographics

For this study cohort, 39 high-risk children for ASD were included in data analysis. The participants included 22 ASD children and 17 non-ASD children. Among them, 25 were males and 14 females; 15 were White (42.8%), 11 Asian (31.4%), and 9 (25.7%) subjects of other races. Table 1 shows the demographic and clinical features of all the participants. There were no significant differences in age or gender between ASD and non-ASD in the two groups; however, their ADOS-2 total and sub-scores were significantly different as expected (Table 1).

**Table 1.** Summary of study participant demographics and ADOS-2 scores.

|  | **ASD [Mean (SD)]** | **Non-ASD [Mean (SD)]** |
| --- | --- | --- |
| Age (years) | 3.8 (1.66) | 3.3 (1.77) |
| **ADOS-2 Scores** |  |  |
| Total | 7.77 (1.8) | 4.47 (2.32) |
| Social Affect | 8 (1.72) | 5.41 (2.9) |
| Restrictive and Repetitive Behavior | 7.09 (1.97) | 3.18 (2.24) |
| **Sex (n)** |  |  |
| Male | 16 | 9 |
| Female | 6 | 8 |
| **Race/Ethnicity (n)** |  |  |
| White | 8 | 9 |
| African American | 2 | 0 |
| Asian | 7 | 6 |
| Hispanic | 3 | 0 |
| Multi-racial | 2 | 2 |

### The Difference of FAS and AVC Between ASD and Non-ASD Children

Video 1 (25 seconds) features a woman (AOI-1 is her face) on the left side of the screen and a tablet (AOI-2) on the right side of the screen. The video is divided into four time blocks (1-2-3-4) as described in the protocol (Figure 1): Block 1 is when the tablet is on with moving pictures, intended to draw the subjects’ attention. Block 2 is when the woman suddenly turns off the tablet, and we expect subjects to turn and look at the woman’s face, wondering what is happening. Block 3 is when the woman turns the tablet on again, and Block 4 is when she turns the tablet off again. Attention shifts were expected during these on-off-on-off transitions of the tablet. The blue bars represent the Total Gaze Count (TGC) of the non-ASD group, and the red bars represent the TGC of the ASD group. Green-colored areas indicate the Favored AOI Shifts (FAS) pathway, representing the expected sequence of attention shifts from tablet to face and back to tablet and face again. Pink-colored areas represent Unfavored Attention Shifts (UAS), as shown in Figure 1.

**Figure 1.**
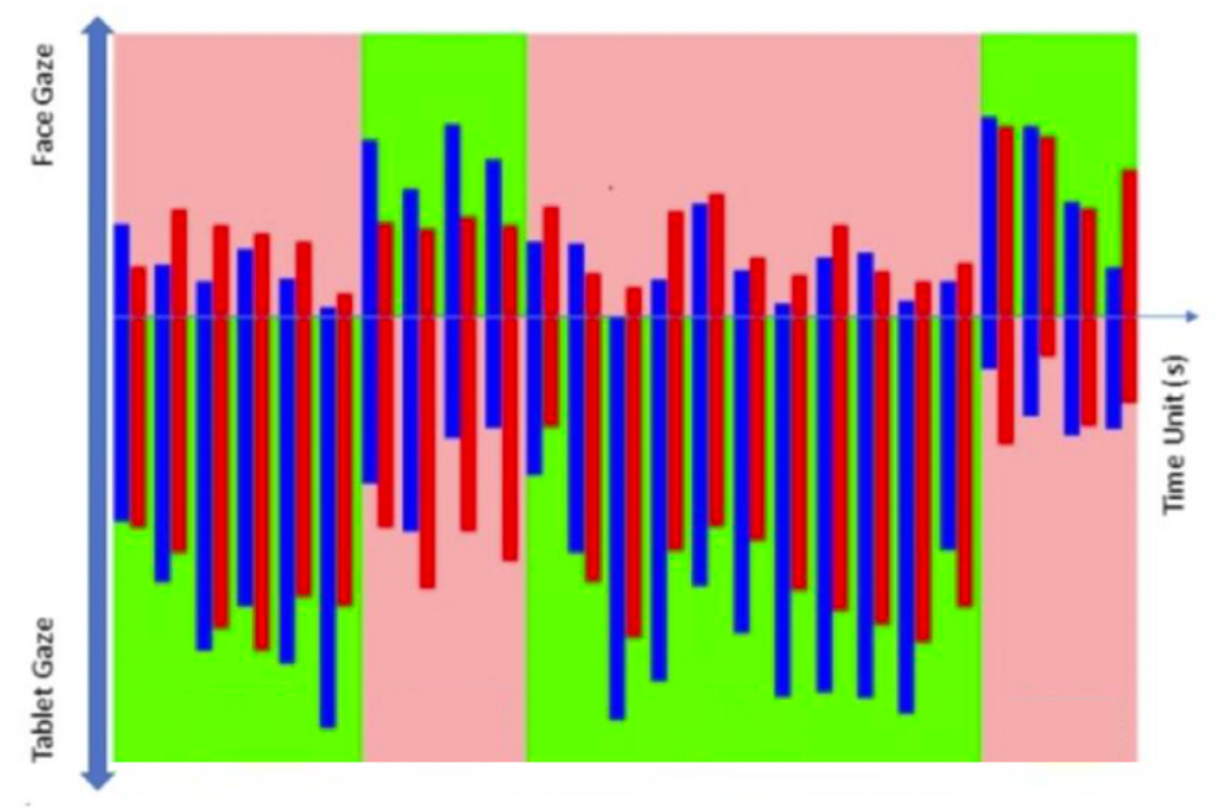
Mean gaze numbers in each second (25s in total) for Video 1, where AOI 1 (face) and AOI 2 (tablet) are shown. Blue bars: non-ASD group; Red bars: ASD group. Green areas: Favored AOI Shift (FAS). Pink areas: Unfavored AOI shift (UAS).

The Total Gaze Count (TGC) was analyzed for both ASD and non-ASD groups across two AOIs during different time blocks (Table 2). We found that non-ASD children showed significant TGC differences across time blocks 1→2, 2→3, and 3→4 for both AOI areas. In contrast, ASD children showed no TGC difference during the 1→2 and 2→3 shifts, and only began to show a difference during the 3→4 shift for both AOI areas. Additionally, the difference between both subject groups was significant during the 1→2 and 2→3 shifts but not during the 3→4 shift (Table 2). When we further investigated the Favored AOI Shifts (FAS) pathway and AOI Vacancy Counts (AVC), which indicate the subject’s gaze counts on neither AOI, we found that the ASD group had significantly reduced TGC along the FAS pathway (p < 0.00001) and significantly increased AVC (p < 0.00001) across different time blocks relative to the non-ASD group (Table 3).

**Table 2.** The comparison of significance of total gaze counts cross time blocks in ASD vs non-ASD groups.

| Time Block | AOI | Intra-time Block *P*-value |  | Groupwise *P*-value |
| --- | --- | --- | --- | --- |
|  |  | Non-ASD | ASD |  |
| 1 → 2 | Face | 0.000011 | 0.8261 | 0.00554 |
|  | Tablet | 0.000055 | 0.1004 | 0.1508 |
| 2 → 3 | Face | < 0.00001 | 0.1491 | 0.01682 |
|  | Tablet | 0.000033 | 0.9966 | 0.01735 |
| 3 → 4 | Face | < 0.00001 | 0.000733 | 0.5203 |
|  | Tablet | < 0.00001 | 0.000039 | 0.1704 |

**Table 3.** The comparison of favored shifts and vacant attentions in ASD vs non-ASD groups.

|  |  | Non-ASD TGC | ASD TGC | *P*-value |
| --- | --- | --- | --- | --- |
| AOI | Face | 314. ± 608 | 288. ± 651 | 0.5249 |
|  | Tablet | 995. ± 1030 | 789. ± 1043 | 0.002142 |
| FAS |  | 1022. ± 1039 | 737. ± 1022 | < 0.00001 |
| FAS-UAS |  | 735. ± 1281 | 396. ± 1341 | 0.00007 |
| AVC |  | 4.53 ± 9.64 | 9.18 ± 12.1 | < 0.00001 |

### The difference in ASC and AVC between ASD and non-ASD children

Video 2 (10 seconds) featured a woman sitting and mouthing the alphabet without sound. We defined two important Areas of Interest (AOIs): AOI-1 (the eye area) and AOI-2 (the mouth area). We studied the Total Gaze Count (TGC) in these AOIs and the AOI Switch Counts (ASC), which represent the switches between these two AOIs. During our analysis, we visualized the ASD and non-ASD groups through the usage of red and blue-colored dots, respectively (figure not shown), for TGC and TFT. The density distribution patterns differ between the ASD and non-ASD groups, with the ASD group displaying a more diverse and scattered distribution.

When we investigated the detailed Total Gaze Count (TGC), AOI Switch Counts (ASC), and AOI Vacancy Counts (AVC), we found that the ASD group had significantly fewer ASCs between AOI 1 and AOI 2 (p = 0.0452) and significantly more AVCs (p = 0.000017) compared to the non-ASD group. Additionally, the TGC was significantly smaller in the ASD group for the AOI-1 area (p = 0.00379) but not for the AOI-2 area (p = 0.6537). In contrast, when comparing the old ET metrics—Total Fixation Time (TFT) and pupil size—there were no significant differences between the two groups (p > 0.05). This demonstrates that AVC and ASC had significantly higher sensitivity than the older metrics (Table 4).

**Table 4.** Comparison of old and new eye tracking metrics in ASD vs non-ASD group for video 2.

|  |  | non-ASD | ASD | P-value |
| --- | --- | --- | --- | --- |
| Eye (AOI-1) | Total gaze count (TGC) | 133.3 | 76.0 | 0.00379 |
|  | Total fixation time (TFT) | 91.24 | 56.96 | 0.0661 |
|  | Pupil Size | 3.316 | 3.456 | 0.7485 |
| Mouth (AOI-2) | Total gaze count (TGC) | 194.5 | 180.4 | 0.6537 |
|  | Total fixation time (TFT) | 163.1 | 159.2 | 0.902 |
|  | Pupil Size (mm) | 3.475 | 3.142 | 0.474 |
| ASC | Between AOIs 1 and 2 | 5.94 | 4.23 | 0.0452 |
| AVC | Total gaze counts | 1.812 | 3.950 | 0.000017 |

### Correlation of significant new ET metrics and ADOS-2 scores/ASD diagnosis

We conducted a correlation study with regression analysis between the significant ET index and ASD severity based on ADOS-2 scores. We found that the Favored AOI Shifts (FAS) to Unfavored AOI Shifts (UAS) ratio for Video 1 negatively correlated with ADOS-2 total scores (r = -0.373, p = 0.01948), Social Affect (SA) scores (r = -0.33, p = 0.0412), and Restricted and Repetitive Behaviors (RRB) scores (r = - 0.25, p = 0.124). Using an ADOS-2 total CSS cut-off score of 5 and a FAS-UAS cut-off score of 641.1, we achieved a sensitivity of 91% and a specificity of 72% (Figure 2).

**Figure 2.**
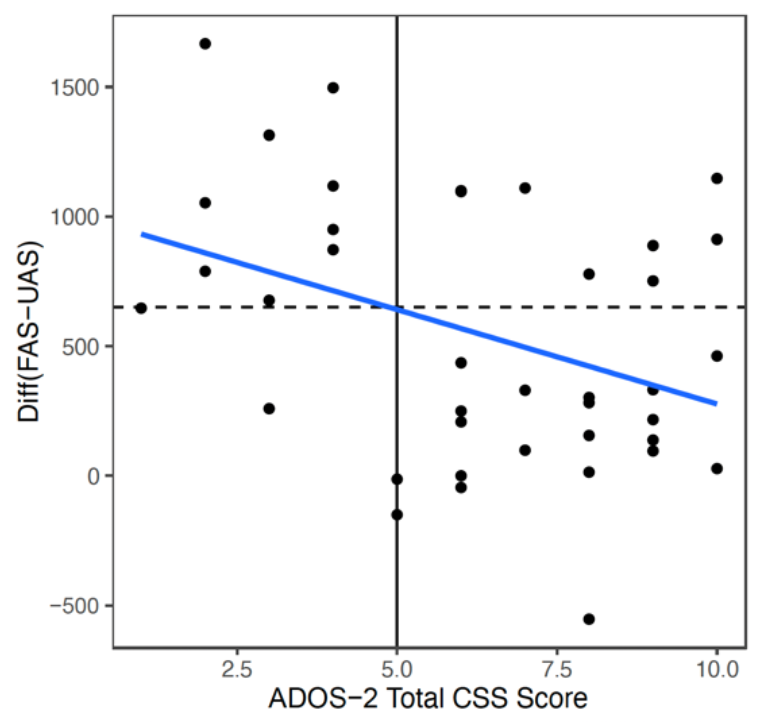
Video 1 (Face and Tablet) FAS-UAS vs ADOS-2 total scores and the linear regression fit. Linear fit: gaze = -72.841*ADOS-2 total scores + 1005.3. Correlation = -0.373. If ADOS-2 total scores cutoff = 5, the gaze cutoff was calculated as: gaze cutoff = 641.1 (below would be diagnosed), specificity = 0.91, sensitivity = 0.72, p = 0.01948.

When comparing the ASD (red dots) versus non-ASD (blue dots) groups, we found that AVC had the highest sensitivity and specificity among all the ET metrics, both new and old. In video 1, AVC demonstrated a sensitivity of 88%, a specificity of 88%, and a p-value of less than 0.00001, with a cut-off score of 0.305 (Figure 3). In video 2, AVC showed a sensitivity of 100%, a specificity of 80%, and a p-value of less than 0.000045, with a cut-off score of 0.306 (Figure 4). Additionally, ASC for video 2 exhibited a sensitivity of 71%, a specificity of 64%, a p-value of 0.04523, and a cut-off score of 4.5.

**Figure 3.**
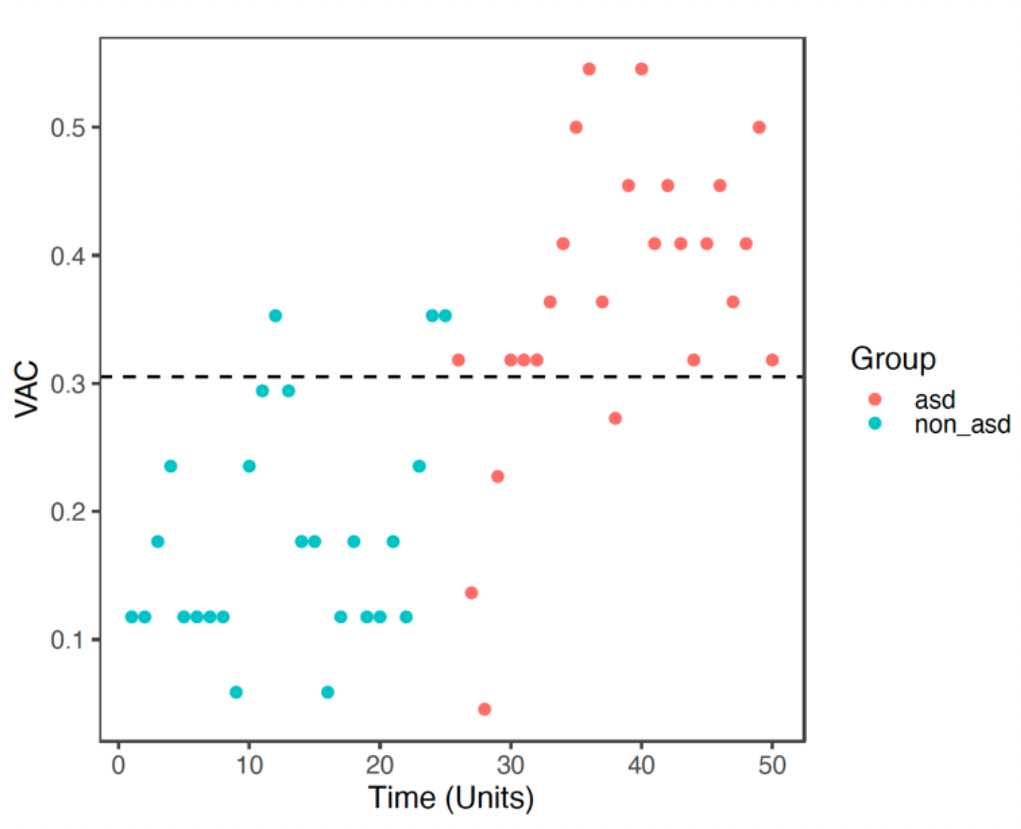
Video 1 (Face and Tablet) VAC vs time unit distributions for non-ASD (round) and ASD (triangle). Results based on the cutoff effect: cutoff = 0.305 (above would be diagnosed), Specificity = 0.88, Sensitivity = 0.88, p < 0.00001.

**Figure 4.**
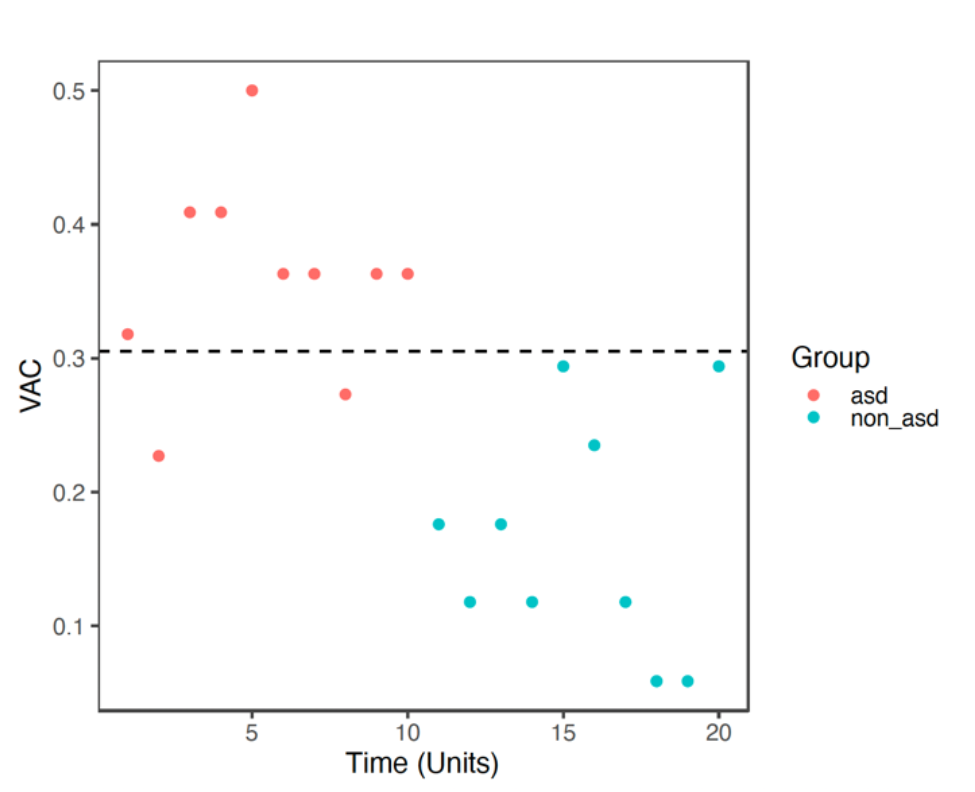
Video 2 (Speaking) VAC vs time unit distributions for non-ASD (round) and ASD (triangle). Results based on the cutoff effect: cutoff = 0.306 (above would be diagnosed), specificity = 1.00, sensitivity = 0.80, p = 0.000045.

## Discussion

Previous investigations into eye tracking (ET) as a diagnostic tool for Autism Spectrum Disorder (ASD) have shown promising results, such as notable reductions in Total Fixation Time (TFT) of Areas of Interest (AOIs) among ASD subjects (Kong, 2022; Papagiannopoulou, 2014). However, the diversity in ET protocols and paradigms has often limited its reliability as a diagnostic instrument (Mastergeorge, 2021).

In response, our study developed and assessed a novel set of ET metrics—Favored AOI Shifts (FAS), AOI Switch Counts (ASC), and AOI Vacancy Counts (AVC)—designed to enhance diagnostic accuracy by capturing subtle behavioral markers fundamental to ASD.

The introduction of FAS versus Unfavored AOI Shifts (UAS) aims to differentiate between typical and atypical attention shifts, reflecting an individual’s ability to dynamically prioritize relevant stimuli. This approach underscores cognitive flexibility, a critical aspect often impaired in ASD. ASC and AVC, respectively, quantify transitions between competitive targets and the absence of gaze on expected targets, providing a nuanced understanding of attentional engagement and disengagement in ASD subjects.

Our findings indicate these new metrics are more sensitive and specific than traditional measures like TFT and pupil size, particularly in distinguishing ASD from non-ASD in a cohort of high-risk toddlers and preschoolers. For instance, our analysis revealed that ASD participants exhibited significantly fewer FAS and heightened UAS during tasks designed to test joint attention (JA), a fundamental social communicative skill that is typically disrupted in ASD (Mundy, 2018). Notably, the correlation of these metrics with ADOS-2 scores suggests that the severity of social affect impairments is inversely related to engagement in favored gaze patterns. The AVC metric emerged as particularly insightful, revealing that ASD subjects frequently failed to engage with designated AOIs—indicative of the inattentive phenomenon often reported anecdotally by caregivers of children with ASD. The statistical robustness of AVC (sensitivity of 88-100% and specificity of 80-88% across various tests) supports its potential utility in clinical settings, emphasizing its role in detecting divergent attention patterns.

This study not only reaffirms the utility of ET in ASD diagnosis but also introduces new avenues for understanding the neural and cognitive underpinnings of the disorder. The reduced tendency of ASD individuals to shift attention as expected may reflect broader deficits in theory of mind and social cognition, potentially linked to underlying neural abnormalities in networks involving the cerebellum and prefrontal cortex (Clausi, 2021; Kelly, 2020). Moreover, while our results are promising, the specificity of the participant cohort, consisting of high-risk individuals rather than a broader demographic including neurotypical controls, necessitates cautious interpretation. Future research should aim to validate these findings across more diverse populations and clinical settings, enhancing the generalizability of the metrics. Our study positions ET not only as a feasible diagnostic tool for early ASD screening but also highlights its potential to provide deeper insights into the distinct neurodevelopmental trajectories associated with the disorder. The development of ET metrics like FAS, ASC, and AVC marks a significant advance in the objective measurement of core ASD features, paving the way for more targeted interventions and therapies tailored to individual neurodevelopmental profiles. As we continue to refine these metrics and explore their implications, larger-scale studies will be essential for establishing their efficacy and integrating them into routine clinical practice.

## Data Availability

All data produced in the present study are available upon reasonable request to the authors.

## List of Abbreviations

ET: Eye-tracking
ASD: Autism Spectrum Disorder
AOI: Area of Interest
ASC: AOI Switch Counts
FAS: Favored AOI Shifts
UAS: Unfavored AOI Shifts
AVC: AOI Vacancy Counts
ADOS-2: Autism Diagnostic Observation Schedule, Second Edition
DSM-5: Diagnostic and Statistical Manual of Mental Disorders, Fifth Edition
TFT: Total Fixation Count
TGC: Total Gaze Count
M-CHAT: Modified Checklist for Autism in Toddlers
SA: Social Affect
RRB: Restrictive and Repetitive Behaviors
CSS: Calibrated Severity Score
ToM: Theory of Mind
ADHD: Attention Deficit and Hyperactive Disorder

## Acknowledgement

We appreciate Dr. Bruce Rosen and Dr. William Stone for their generous support and advise, we thank Dr. Xiaochun Wang for his comments and assistance for the manuscript, and we thank Zexu Li for his meticulous effort in manuscript editing and formatting. We thank all the participants and their parents for the great contribution for this work.

## Ethics and Informed Consent

The study was conducted according to the guidelines of the Declaration of Helsinki and approved by the Institutional Review Board of Massachusetts General Hospital 2017P001667, 13 July 2018). The secondary use of research samples/data was approved by the Institutional Review Board of Massachusetts General Hospital (2020P004102; January 7, 2021). Informed consent was obtained from all subjects involved in the study.

## Consent for Publication

All authors agree with the publication of the current manuscript.

## Data Availability Statement

The data presented in this study are available on request from the corresponding author.

## Competing Interests

The authors declare that the research was conducted in the absence of any commercial or financial relationships that could be construed as a potential conflict of interest.

## Funding

This research was funded by Massachusetts General Hospital, grant numbers #230361 and #233263.

## Author Contributions

Conceptualization, X.-J.K.; methodology, RKW, X.-J.K.; software, RKW; validation, RKW; formal analysis, RKW; investigation, X.-J.K, RKW; resources, X.-J.K.; data curation, RKW; writing—original draft preparation, RKW, writing, review and editing, RKW, X.-J.K, KL,KK; visualization, RKW, KL; supervision, X.-J.K.; KK, project administration, X.-J.K. KK; funding acquisition, X.-J.K. All authors have read and agreed to the published version of the manuscript.

## Notes

### Competing Interest Statement

The authors have declared no competing interest.

### Funding Statement

This study was funded by Massachusetts General Hospital, grant numbers #230361 and #233263.

### Author Declarations

Institutional Review Board of Massachusetts General Hospital gave ethical approval for this work. (2017P001667, 13 July 2018; 2020P004102; January 7, 2021)

